# Risk Factors for COVID-19 versus non-COVID-19 related in-hospital and community deaths by Local Authority District in Great Britain

**DOI:** 10.1101/2020.05.21.20108936

**Authors:** Samuel P Leighton, Danielle J Leighton, James Herron, Rachel Upthegrove, Jonathan Cavanagh, Georgios Gkoutos, Breda Cullen, Pavan K Mallikarjun

## Abstract

**Objectives:** To undertake a preliminary hypothesis-generating analysis exploring putative risk factors for coronavirus diseae 2019 (COVID-19) population-adjusted deaths, compared with non-COVID-19 related deaths, at a local authority district (LAD) level in hospital, care homes and at home.

**Design:** Ecological retrospective cohort study

**Setting:** Local authority districts (LADs) in England, Scotland and Wales (Great Britain (GB)).

**Participants:** All LAD deaths registered by week 16 of 2020.

**Main Outcome Measures:** Death registration where COVID-19 is mentioned as a contributing factor per 100,000 people in all settings, and in i) cares homes, ii) hospitals or iii) home only, in comparison to non-COVID-19 related deaths.

**Results:** Across GB by week 16 of 2020, 20,684 deaths had been registered mentioning COVID-19, equivalent to 25.6 per 100,000 people. Significant risk factors for LAD COVID-19 death in comparison to non-COVID-19 related death were air pollution and proportion of the population who were female. Significant protective factors were higher air temperature and proportion of the population who were ex-smokers. Conversely, for all COVID-19 unrelated deaths in comparison to COVID-19 deaths, higher rates of communal living, higher population rates of chronic kidney disease, chronic obstructive pulmonary disease, cerebrovascular disease deaths under 75 and dementia were predictive of death, whereas, higher rates of flight passengers was protective. Looking at individual setttings, the most notable findings in care homes was Scotland being a significant risk factor for COVID-19 related deaths compared to England. For hospital setting, the proportion of the population who were from black and Asian minority ethnic (BAME) groups significantly predicted COVID-19 related death.

**Conclusions:** This is the first study within GB to assess COVID-19 related deaths in comparison to COVID-19 unrelated deaths across hospital, care homes and home combined. As an ecological study, the results cannot be directly extrapolated to individuals. However, the analysis may be informative for public health policy and protective measures. From our hypothesis-generating analysis, we propose that air pollution is a significant risk factor and high temperature a significant protective factor for COVID-19 related deaths. These factors cannot readily be modelled at an individual level. Scottish local authorities and local authorities with a higher proportion of individuals of BAME origin are potential risk factors for COVID-19 related deaths in care homes and in hospitals, respectively. Altogether, this analysis shows the benefits of access to high quality open data for public information, public health policy and further research.

## Introduction

The global coronavirus disease 2019 (COVID-19) crisis has presented the United Kingdom (UK) with an unprecedented challenge in the management and prediction of infection and mortality rates. With the benefit of frequently updated open-access population health data, it is possible for researchers in the UK to identify regional influences of disease progression and outcome that, crucially, might help to model service provision in real-time.

Studies of disease trajectory related to COVID-19 have emerged over the past few weeks and have highlighted several demographic and clinical factors that influence outcome. Initial data from Wuhan suggested that increased age and presence of co-morbidities, in particular cardiovascular disease, predicted death (as compared with discharge) in a cohort of patients admitted to hospital with Severe Acute Respiratory Syndrome Coronavirus 2 (SARS-CoV-2) infection.[1,2] A meta-analysis of 44,672 laboratory-confirmed cases in China confirmed these findings, and also highlighted the increased risk of death among men, with an equal rate of infection among males and females.[3]

In a series of 4103 patients in New York City, 48.7% were hospitalised, with age >65 and obesity most strongly predicting admission; tobacco use was an independent negative predictor of hospitalisation.[4] A further multivariable report of 16,749 patients from 166 UK hospitals (33% death rate) confirmed that increased age was strongly independently associated with death and that co-morbidities (in order of importance: dementia, obesity, chronic cardiac disease, chronic kidney disease (CKD), chronic obstructive pulmonary disease (COPD) and malignancy) were also independently associated with mortality.[5] Females were 20% less likely to die than males. However, the study did not control for ethnicity, smoking status, local population factors such as social deprivation, population density, population and goods movement, or ecological factors such as climate and pollution.

Risk factors for in-hospital deaths have also been explored using a large cohort of primary care patients in England[6]. Primary care linkage data permitted detailed analysis of past medical and demographic risk factors, which were studied in individual age and sex-adjusted models; significant variables were then incorporated into a multivariable model. Again, co-morbidities were considered to impart significant risk of death, as well as deprivation and being from Asian and black ethnicity minorities. Current smoking was found to be protective against death from COVID-19 but lost significance when adjusted for ethnicity.

Ethnicity has recently been identified as a research priority, following observations from the Institute of Fiscal Studies that COVID-19 related death rates of people of black, Asian and minority ethnic (BAME) origin are 2.5 times that of people of white ethnic origin in English hospitals. Exclusion of ethnicity data from some multivariable analyses thus far is a concerning omission.[7]

Ecological modelling suggests that ‘lived’, or perceived, population density, and climate are thought to modify the rate of transmission of SARS-CoV-2.[8,9] Moreover, direct links between air pollution and the severity of respiratory disease or inflammatory response in COVID-19 disease have been postulated in Northern Italy,[10] England[11] and the United States.[12] Cultural and behavioural variables are more difficult to study but, inevitably, local, domestic and international travel and transport[13,14] and public and cultural awareness[15] is thought to contribute to population spreading.

While respiratory failure is the most common cause of death of people with COVID-19 in hospital[1], evidence for an overwhelming burden of out-of-hospital deaths has emerged in the UK. Possible reasons for this include: i) recent proactivity from primary care providers in formalising ‘Do Not Attempt Resuscitation’ orders in care home residents and individuals over 70 with co-morbidities; ii) triaging of intensive care admissions; iii) expected deaths in those with co-morbidities with concomitant infection with SARS-CoV-2 being an “epiphenomenon”.[16] However, other independent disease-specific or location-specific influences may contribute. Systematic analysis of risk factors for COVID-19 related deaths in the community is currently lacking.

Death rates in Great Britain are anticipated to be amongst the highest in Europe,[17] although variation in reporting of out-of-hospital deaths currently makes between-country comparisons difficult.[18] In light of ongoing fatalities, factors influencing survival may have broader public health implications and help to model services at a local authority level. Using publicly available aggregate Local Authority District (LAD) data, we undertook a multivariable, multivariate ecological study of SARS-CoV-2 infection related deaths in England, Scotland and Wales in hospitals, care homes and at home. We anticipated that these data would: i) generate further data-driven hypotheses regarding risk factors for death related to COVID-19 at a local authority level; ii) identify differences between risk factors for COVID-19 related deaths and deaths due to all other causes; and iii) identify risk factors specific to hospital, care home and at home COVID-19 related deaths.

## Aim

To undertake a preliminary hypothesis-generating analysis to explore putative risk factors for novel coronavirus (COVID-19) population-adjusted death rates, compared with non-COVID-19 related deaths, at a local authority district (LAD) level in hospital, care homes and at home.

## Methods

We adhered to the Strengthening the reporting of observational studies in epidemiology (STROBE) guidelines for the reporting of this observational study.[19]

### Sources of data

All data used in the present analysis is free, publicly available, and open source.

#### Outcome

We used open-source government data for total English, Scottish and Welsh (hereafter referred to as Great Britain (GB)) death registrations mentioning COVID-19 by LAD for week 16 obtained from the Office for National Statistics (ONS) for England and Wales (registrations to 17^th^ April 2020), and from the National Records of Scotland (NRS) for Scotland (registrations to 19^th^ April 2020).[20,21] The International Classification of Diseases, 10^th^ Edition (ICD-10) definitions are as follows: COVID-19 (U07.1 and U07.2).[22] At the time of analysis, Northern Ireland COVID-19 death registrations were not available at a LAD level. Deaths rates were adjusted for GB LAD population obtained from the ONS, per 100,000.[23] GB COVD-19 deaths per 100,000 by LAD were analysed altogether and separately by location – in hospital, care homes and at home. All LAD deaths (except COVID-19) were also analysed across the four settings by way of comparison.

Local authority district (LAD) level is the smallest division UK geographical area for which COVID-19 death rates are publicly released. There are 317 LADs in England made up of 36 metropolitan boroughs, 32 London boroughs, 192 non-metropolitan districts, and 55 unitary authorities, as well as the City of London and the Isles of Scilly. Scotland has 32 LADs equivalent to council areas, and Wales has 22 LADs known as principal areas. In total, there are 371 GB LADs.

#### Putative Risk Factors

Twenty-six putative risk factors for COVID-19 at a LAD level, obtained by literature search and by applying expert clinical knowledge, were included in the present analysis. These relate to country, weather and air pollution; demographics and social influences; population disease rates and behaviours; transport and information behaviour. After dummy coding, this resulted in 27 variables. A literature search was conducted between 28^th^ April and 4^th^ May 2020 independently by SPL, DJL and PKM via Pubmed, Google and Twitter (given the very preliminary nature of the literature surrounding COVID-19 which is mainly available via expert opinion, government briefings and preprint servers), using the search terms “coronavirus” OR “COVID-19” AND “risk” or “risk factors”.

##### Country, Weather and Air Pollution

The constitutive nation of GB that each LAD formed part of was included to reflect the devolved nature of health and social care.

Weather data for average daily maximum temperature and humidity by local authority was obtained from the Met Office[24] using coordinates provided the ONS Open Geography Portal[25] via the Dark Sky application programming interface (API) by Apple.[26] Data was averaged across daily data for four Sundays, 23^rd^ February, 1^st^, 8^th^ and 15^th^ March 2020, because of limitations on the number of free API calls.

Air pollution data for population-weighted LAD 2018 mean total fine particulate matter (PM_2_._5_) was obtained from the UK Government Department for Environment, Food and Rural Affairs.[27]

##### Demographics and Social Influences

Median age, percentage of the population who are female, and population density (people per square kilometre) were obtained from ONS LAD population data.[23] The proportion of LAD population living in a communal establishment (including care homes, prisons, defence bases, boarding schools and student halls), proportion black, Asian and minority ethnic (including Jewish) people (BAME) and proportion living in households with more than 1.5 people per room was obtained from 2011 Census data from the ONS for England and Wales,[28] and NRS for Scotland.[29] Deprivation level for each LAD was defined as the proportion of the LAD’s lower super output areas (LSOAs) in the most deprived 10% of all the constituent nation’s LADs using summary data provided by the 2019 English (EIMD2019), 2020 Scottish (SIMD2020) and the Welsh (WIMD2019) indices of multiple deprivation.[30–32] The proportion of care home residents self-funding – as a surrogate marker for community wealth – was obtained for England regions in 2017 via a House of Commons Library Briefing Paper and mapped to their LADs,[33] for Scottish LADs via the Care Home Census for Adults in Scotland (2017),[34] and for Wales via a 2015 Public Policy Institute for Wales report.[35]

##### Population Disease Rates and Behaviours

For all health and illness LAD authority data, all LAD population prevalences were standardised within the constitutive GB nation, to account for differences in methodology and time of their measurement. Where data for a LAD was not available, health board (Scotland and Wales) or county (England) level data was mapped to LAD. Cancer rates were obtained for England as ‘incidences of all cancers, standardised incidence ratio 2012 – 16’ via Public Health England (PHE),[36] for Scotland as Quality Outcome Framework (QOF) 2015 prevalence via gpcontract.co.uk,[37] and for Wales as QOF 2019 prevalence via StatsWales.[38] Chronic Obstructive Pulmonary Disease (COPD) rates were obtained for England as ‘estimated prevalence of COPD (all ages) 2015’ via PHE, and for Scotland and Wales via QOF, as above. For Chronic Kidney Disease (CKD) rates were obtained for England as ‘CKD prevalence estimates for local and regional populations – 2015’ via PHE, and for Scotland and Wales via QOF. For cardiovascular disease (CVD) age-standardised death rates per 100,000 for coronary heart disease under 75s by LAD 2015/17 were obtained for all GB LADs via the British Heart Foundation.[39] Diabetes rates were obtained for England as ‘estimated prevalence of diabetes (undiagnosed and diagnosed) 2015’, and for Scotland and Wales via QOF. Dementia rates were obtained for England as ‘dementia: QOF prevalence (all ages) 2018/19’ via PHE, and for Scotland and Wales via QOF. Hypertension rates were obtained for England as ‘estimated prevalence of diagnosed hypertension (16+) 2015’ via PHE, and for Scotland and Wales via QOF. Obesity rates for England were obtained as ‘Obesity: QOF prevalence (18+) 2018/19’ via PHE, for Scotland via the Scottish Health Survey 2018,[40] and for Wales via the Welsh Health Survey 2015.[41] Rheumatoid Arthritis (RA) rates were obtained for England as ‘rheumatoid arthritis: QOF prevalence (16+) 2018/19’ via PHE, and for Scotland and Wales via QOF. Finally, current and ex-smoking rates were obtained for England via the GP Patient Survey on PHE, and for Scotland and Wales via their respective 2018 and 2015 health surveys.

##### Transport and Information Behaviour

Population standardised flight passengers to and from each LAD were obtained via the Civil Aviation Authority 2018 Passenger Survey, rounding down to the nearest 100,000 passengers per 100,000 people. Where necessary, county-level figures were divided across their constituent LADs according to their relative populations.[42] Port activity statistics in tonnage were obtained from the UK Government’s Department for Transport and mapped to the port’s LAD for all Major Ports with an activity level greater than the largest Minor Port.[43] Lastly, Google search trend data – as a proxy marker for information-seeking behaviour and public awareness – were obtained from Google Trends for relative search volume for the term “Coronavirus” scaled by population per region (cities, towns and areas of London) mapped to LAD, averaged between 1^st^ February to 19^th^ March 2020.[44]

### Sample Size Calculation

Using Green’s method to calculate sample size for multiple ordinary least squares regression, we would require a minimum of 266 observations for 27 variables (50 + 8 × number of variables).[45] We had outcome data for all 371 GB LADs, exceeding Green’s criteria. Alternatively, using G*Power,[46] at alpha 0.05 and power 0.80, the sample size and number of predictors is sufficient to detect an overall model effect size of f^2^ = 0.07 (equivalent to R^2^ = 0.07).

### Statistical Analysis

#### Univariate Analysis

Country was dummy coded with England as the reference group. Independent variables were standardised to Z-scores. Missing data were handled by multiple imputation (chained equations using predictive mean matching with five nearest neighbours; five imputations) and estimates were pooled using Rubin’s rules.[47] Pearson’s correlation was assessed across the independent variables. Separate multivariable linear regression models with the same independent variables were fitted by ordinary least squares against two sets (COVID-19 deaths and all deaths except COVID-19) of four different dependent variables: LAD deaths per 100,000 people in all settings combined, care homes only, hospitals only, and, at home only. Coefficient estimates with 95% confidence intervals are reported, along with the model’s adjusted R^2^. Assumptions of homoscedasticity and normality of residuals were checked for each outcome model and found not to be violated. We also conducted a sensitivity analysis with robust standard errors which did not change the interpretation of the results.

#### Multivariate Analysis

To establish whether independent variables which were statistically significant in explaining COVID-19 LAD deaths in the univariate multiple regression were the same or different to independent variables which explained all LAD deaths (except those caused by COVID-19), a multivariate multivariable analyses (otherwise known as seemingly unrelated regression) was undertaken within each of the four settings. That is, for each multivariate analysis, two dependent variables, LAD COVID-19 deaths per 100,000 and all LAD deaths (except COVID-19) per 100,000, were regressed against the same 27 standardised independent variables. A type II multivariate analysis of variance (MANOVA) was conducted to assess if the independent variables’ coefficients remained significant in the multivariate multiple regression model via Pillai’s trace. Then, the equality of each multivariate-significant independent variable’s coefficient across the two dependent outcomes was tested using a Wald test, with the null-hypothesis being that the coefficient magnitude was equal for both dependent outcomes (i.e. two-tailed). Significance for the Wald tests was corrected for multiple comparisons by controlling for the false discovery rate. The dependent variables were standardised before performing the Wald tests, to enable a like-for-like comparison across coefficients, given that the outcome prevalence was much larger for deaths except COVID-19 than for COVID-19 deaths. As pooling across the five imputed datasets was not possible for the MANOVA analysis, this analysis and the Wald tests were based on the first imputed dataset only. Replicating the Wald tests with pooling across the five imputed datasets did not alter the interpretation.

All analyses were performed using R CRAN version 3.6.1[48] (with the ‘broom’,[49] ‘car’,[50] ‘mice’,[51] ‘multcomp’,[52] ‘systemfit’[53] and ‘VIM’[54] packages). R code and pre-processed data are available <u>online</u>. Raw data is available from the referenced sources.

### Patient and Public Involvement

No patients were involved in setting the research question or the outcome measures, nor were they involved in the study design. No patients were involved in the interpretation or writing up of results. Results will be disseminated to the public via our respective universities.

## Results

All LADs had complete outcome data. Eighteen of the independent variables had missing data with obesity having the highest percentage missing at 6.5%. Summary statistics for the variables chosen are openly available as referenced above. Across GB by week 16, there had been a total of 20,684 deaths registered mentioning COVID-19 including 3,627 in care homes, 15,668 in hospitals and 1,050 at home. **Figure 1** shows the population-weighted deaths per 100,000 for COVID-19 by week 16 for all settings across each LAD. Of the 371 GB LADs, Inverclyde had the highest reported COVID-19 deaths per 100,000 across all settings at 97.2, while only the Isles of Scilly had no reported COVID-19 deaths. **Supplementary Figure 1** shows the population-weighted deaths per 100,000 for all causes except COVID-19. **Table 1** summarises these numbers. **Figure 2** shows the univariate correlation matrix for the putative risk factor variables.

**Figure 1:**
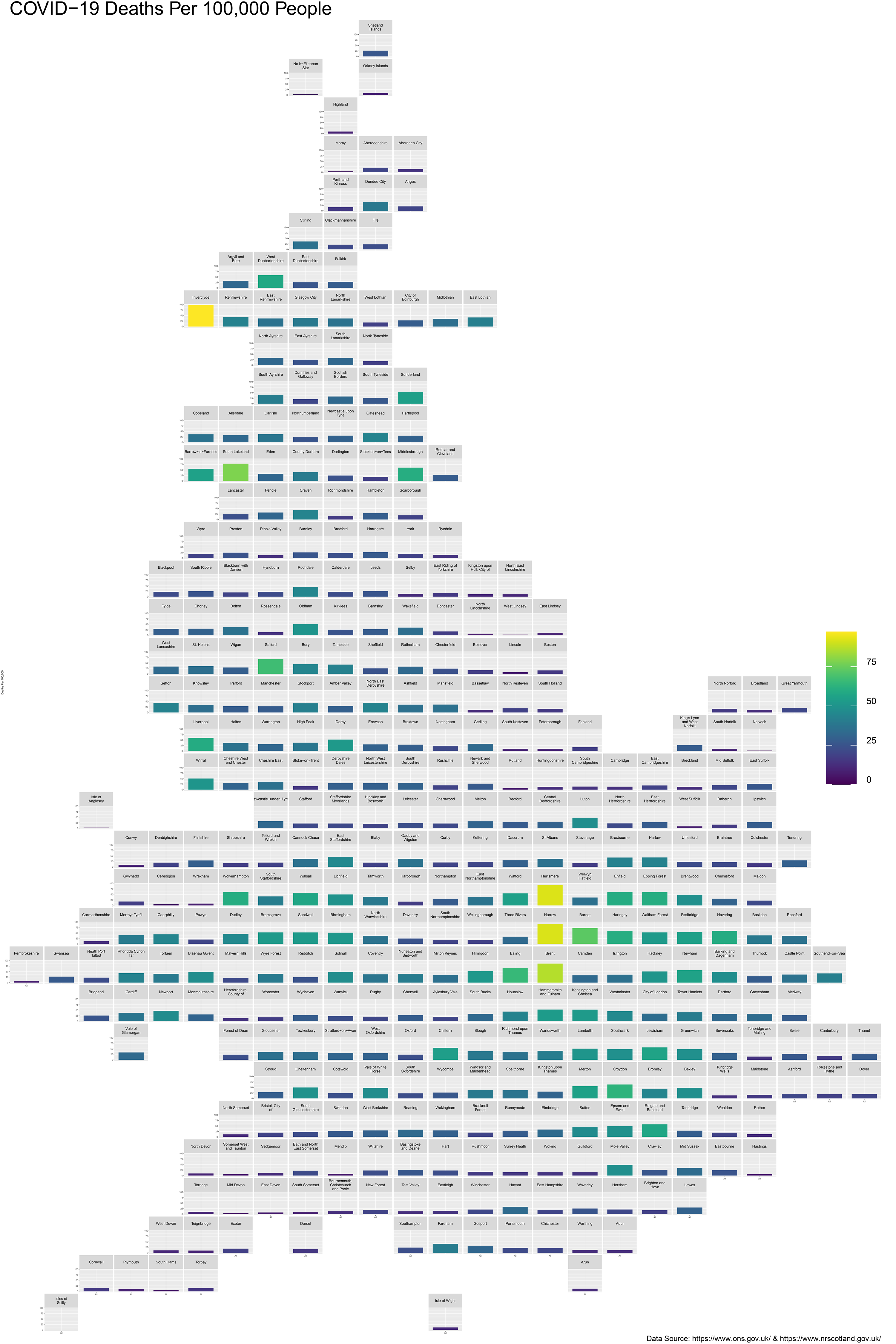
Local Authority District (LAD) COVID-19 deaths per 100,000 people across Great Britian (GB). Data sources: Office for National Statistics (ONS) and National Records of Scotland (NRS).

**Table 1:**
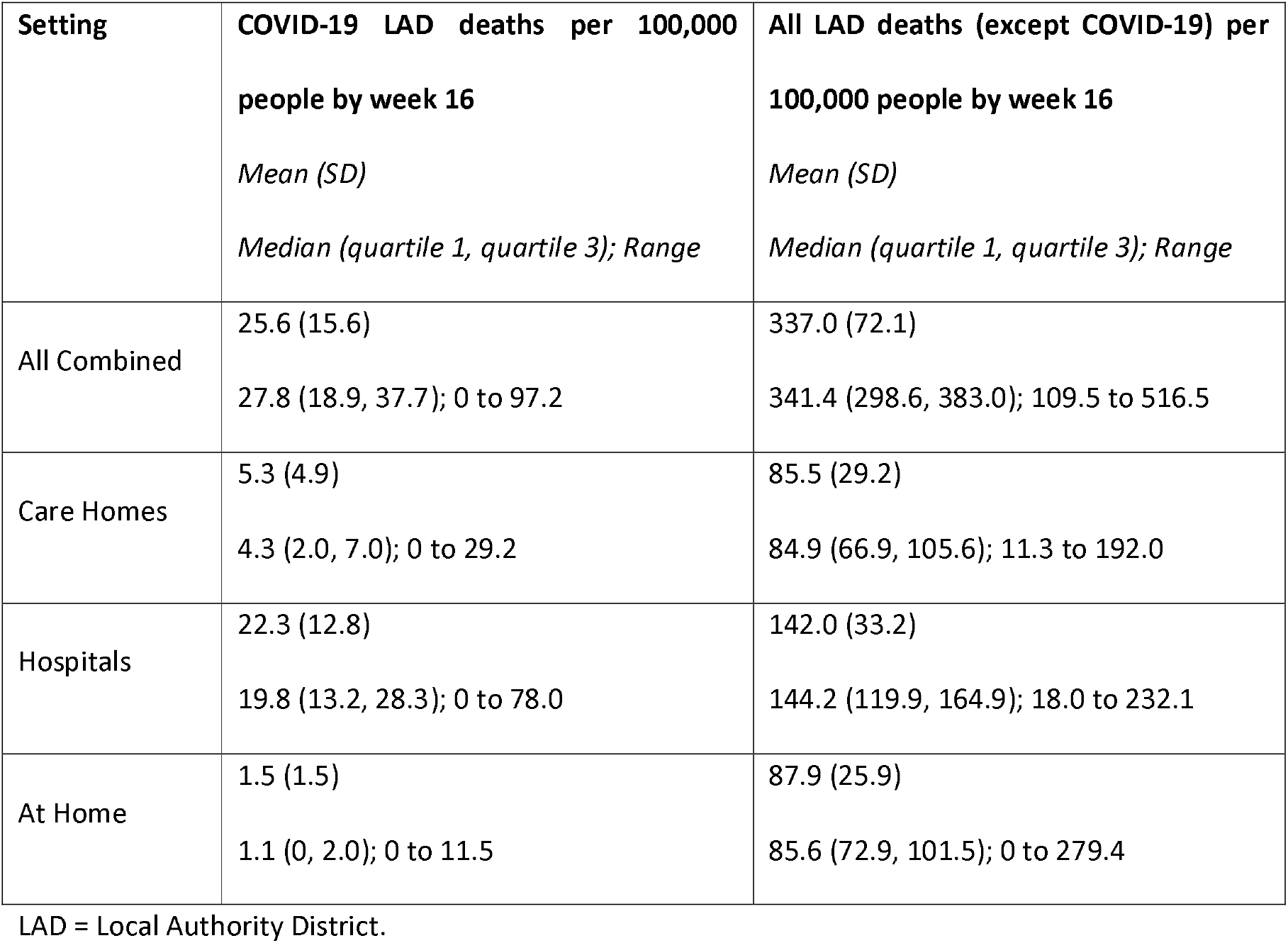
Summary of Death Rates Across Each Setting

**Figure 2:**
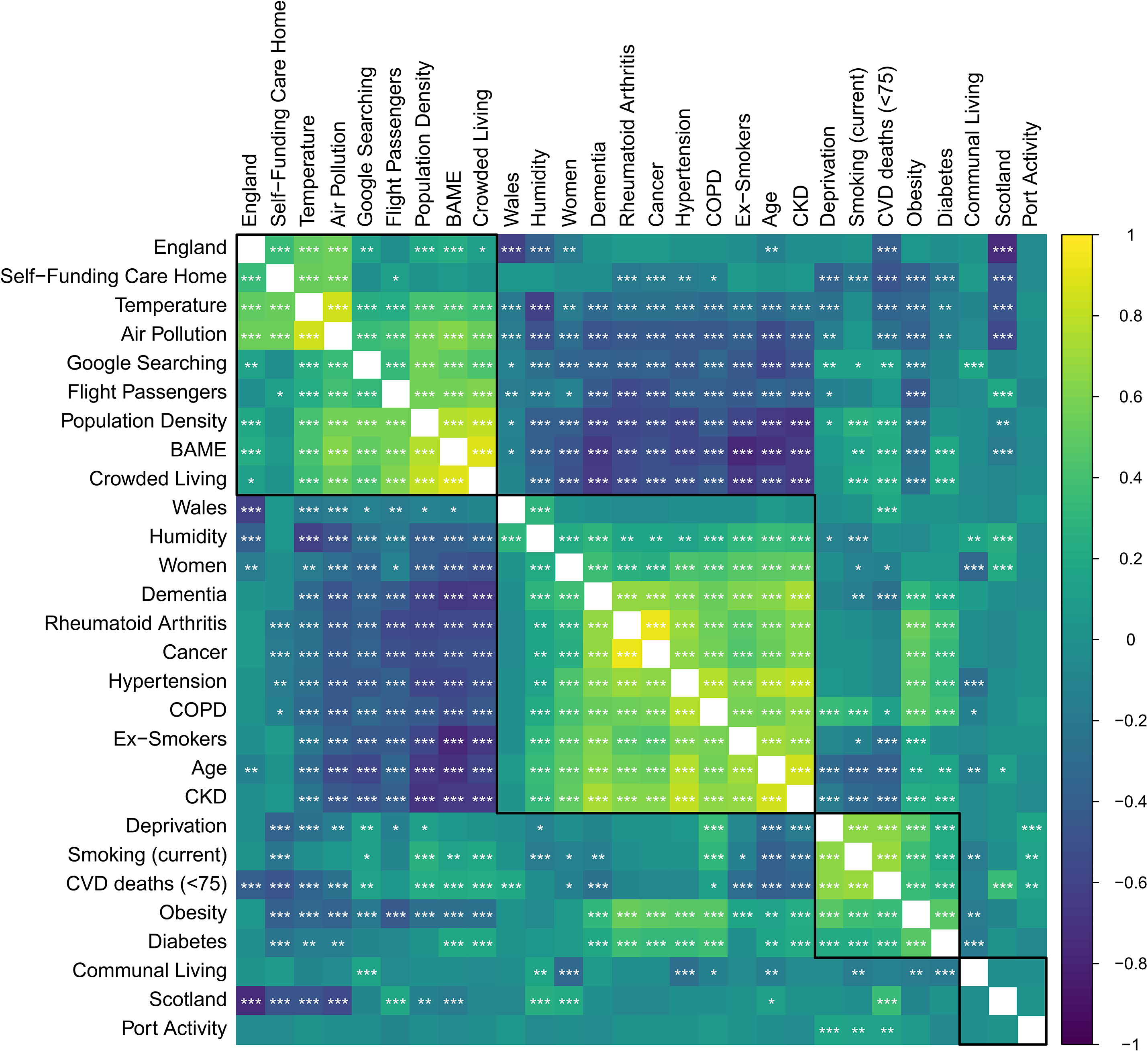
Correlation heatmap (Pearson’s) of putative Local authority risk factors for COVID-19 related deaths. Significance levels: * 0.05-0.01, ** <0.01-0.001, ***<0.001.

**Figure 3** summarises the results of the univariate multiple linear regression analyses for all settings for LAD COVID-19 deaths per 100,000 **(A)** and all LAD deaths (except COVID-19) **(B)**. Significant predictors for COVID-19 related deaths at a local authority level included air pollution, deprivation, and proportion of the population who are female. In contrast, higher temperatures, dementia, exsmokers and self-funding care were significantly protective factors against COVID-19 deaths. The adjusted R^2^ was 0.46 (95% CI 0.38, 0.53), with the model explaining 46% of the variance in the outcome. **Supplementary Figures 2-4** summarise the same analyses for care homes only, hospitals only and at home only, respectively.

**Table 2** shows the independent variables that were significant in the multivariable multivariate analysis of both dependent death outcomes (COVID-19 related versus all except COVID-19 related death rates) across all settings, along with the results of their Wald equality tests. These results highlight significant differences between COVID-19 related and COVID-19 unrelated deaths after correcting for multiple comparisons (†). The Wald test z-value shows the direction of significance (positive value indicating that the risk factor is more strongly associated with COVID-19 related deaths over COVID-19 unrelated deaths). Interpreting these data in the context of **Figures 3A** and **3B**, we can conclude that air pollution (p = 0.007) and higher female population proportion (p = 0.036) are significant risk factors for COVID-19 related deaths at a local authority level compared with all deaths except those related to COVID-19. In contrast, higher temperatures (p = 0.004) and a higher proportion of ex-smokers (p = 0.002) are protective. Conversely, older median age (p = 0.007), higher rates of communal living (p <0.001), higher rates of CKD (p = 0.017), COPD (p = 0.003), CVD deaths (p <0.001) and dementia (p <0.001) are more significant risk factors for deaths that are unrelated to COVID-19 at a local authority level, whereas a higher number of flight passengers (p <0.001) is relatively protective. Female population proportion was also a risk factor for COVID-19 unrelated death, but there was a significantly stronger association in the COVID-19 related death model.

**Table 2:**
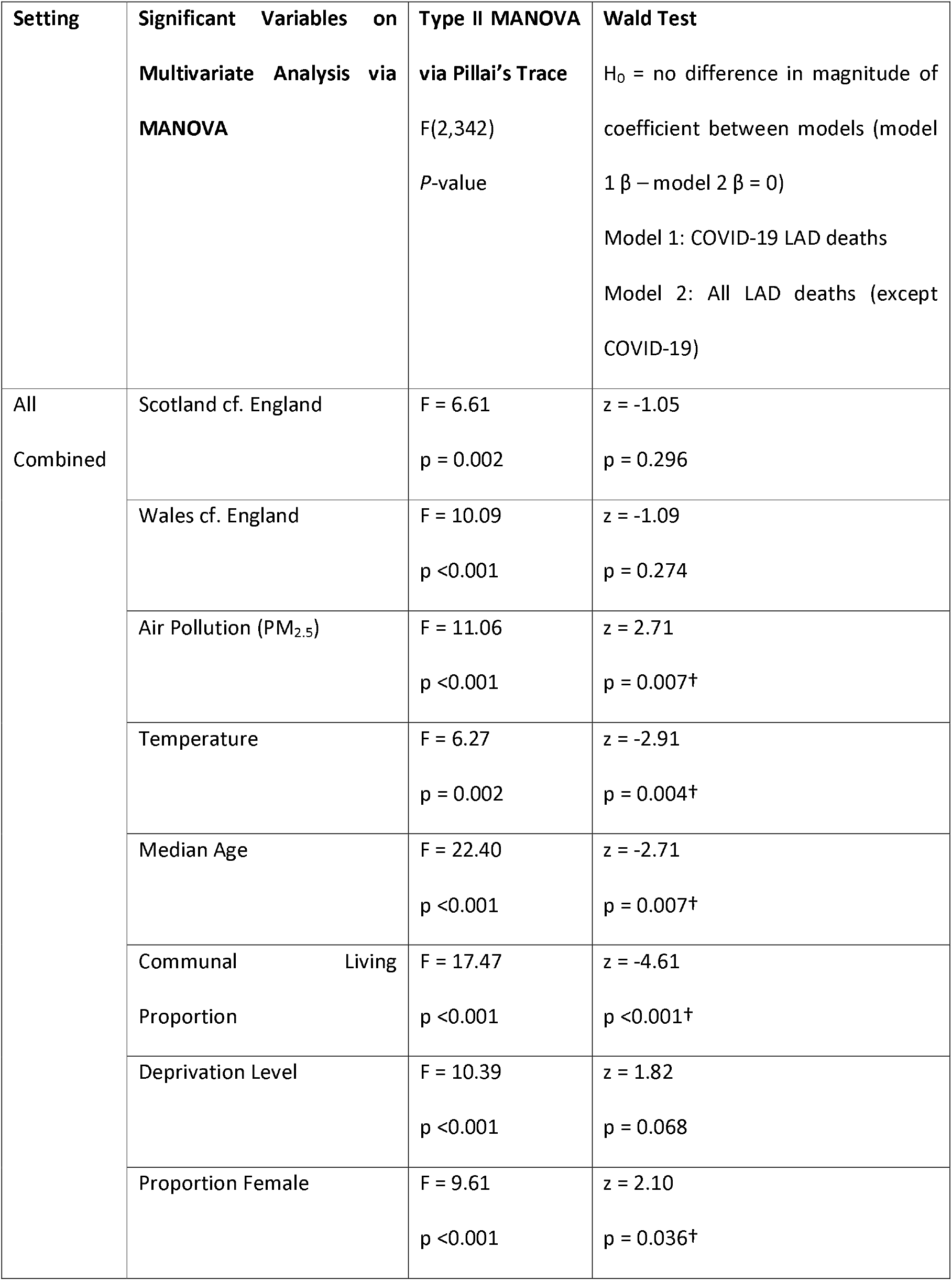
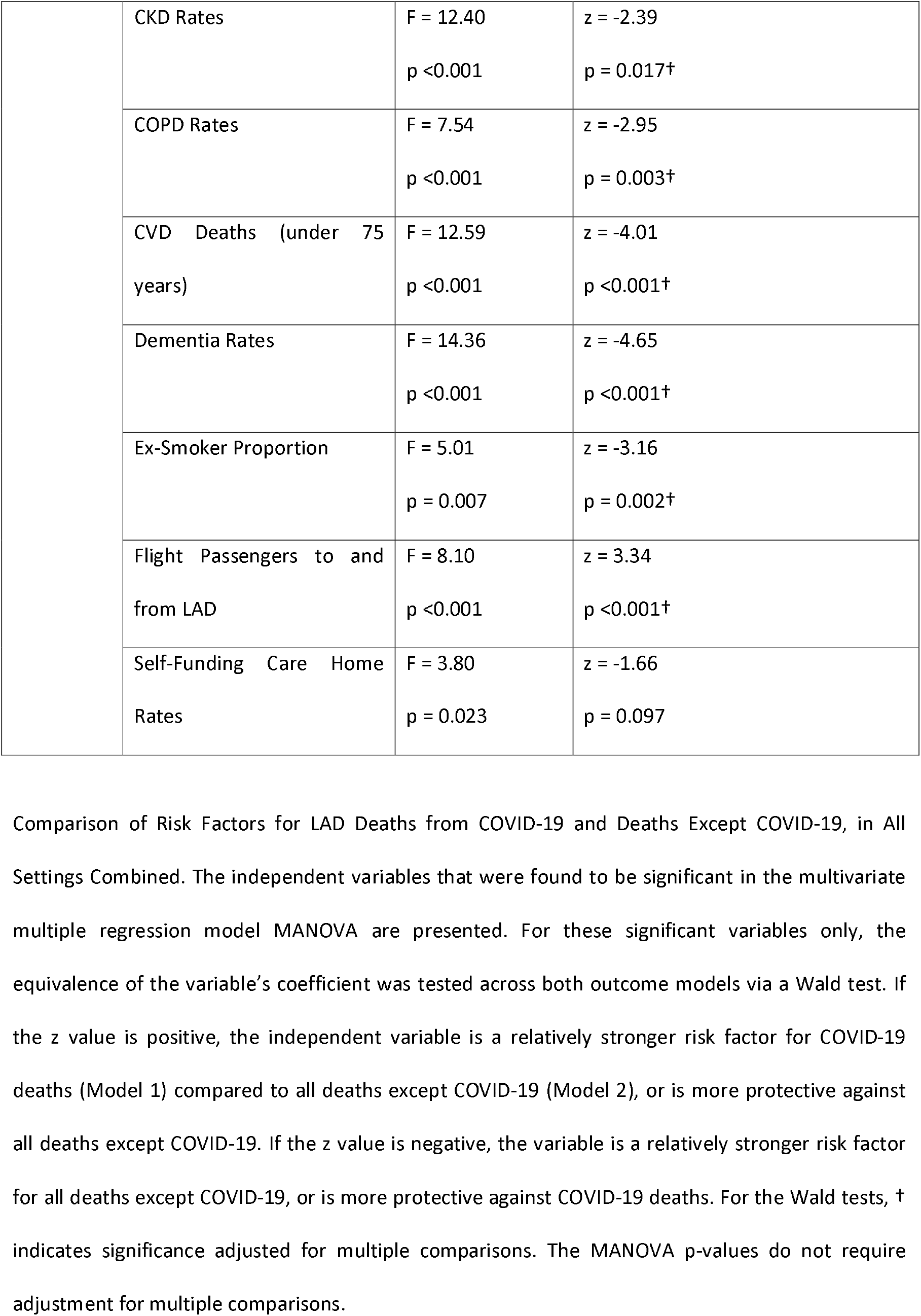
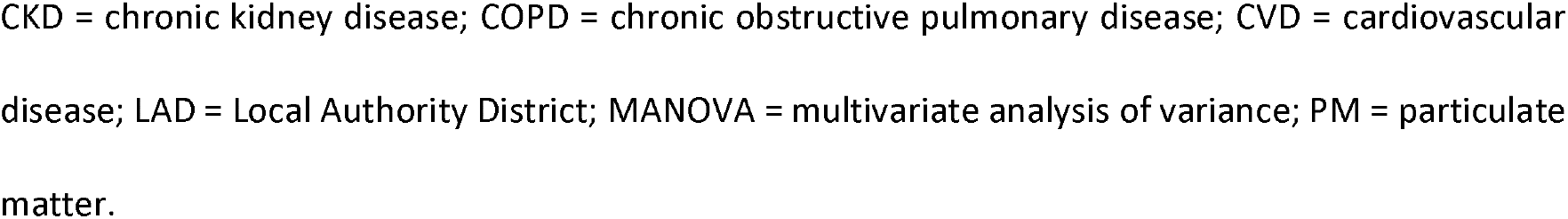
Comparison of Risk Factors for LAD Deaths from COVID-19 and Deaths Except COVID-19, in All Settings Combined

**Figure 3A:**
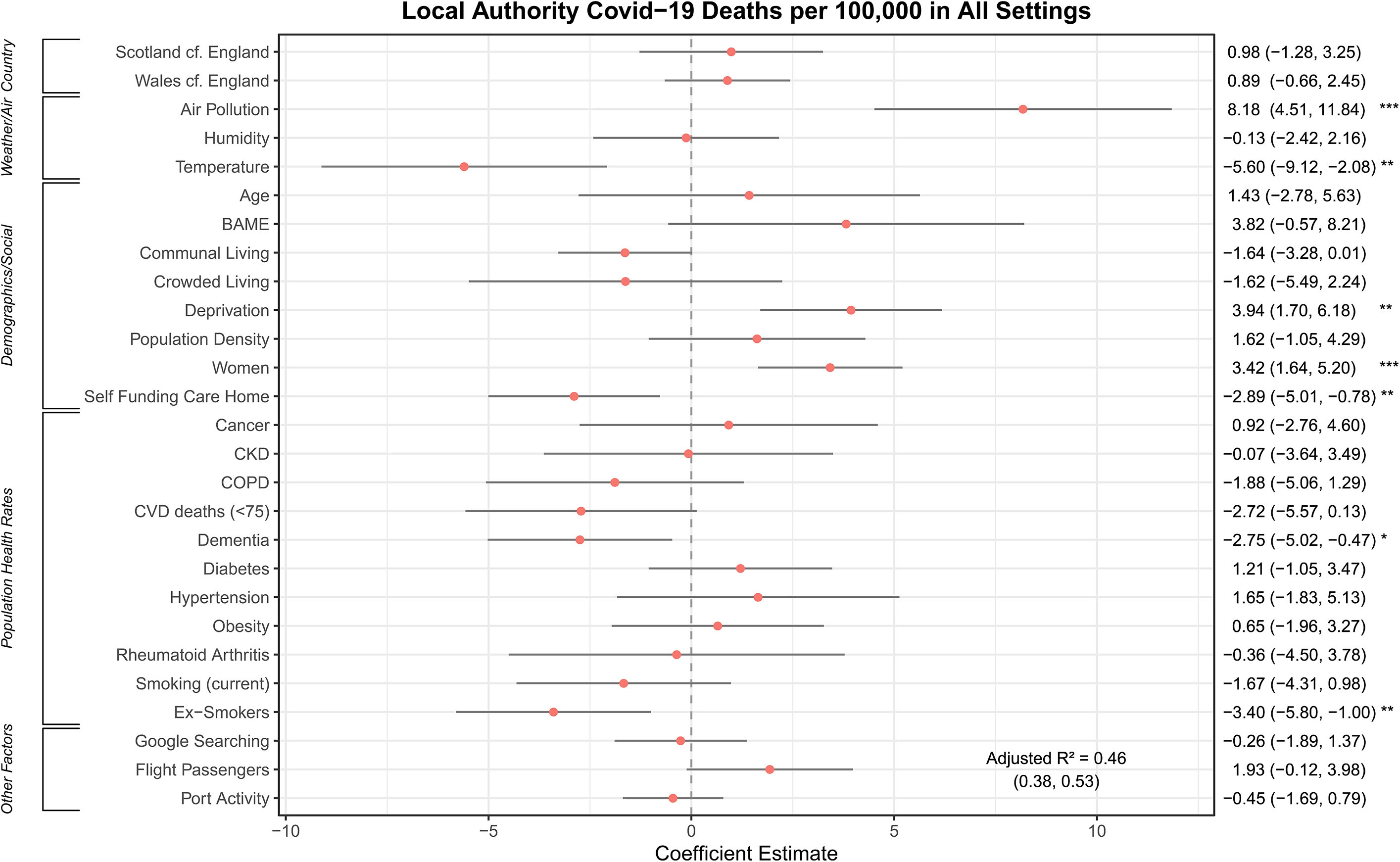
Forest plot of standardised regression coefficients for Local Authority District (LAD) COVID-19 deaths per 100,000 of the population in all settings (care home, hospital and home) with 95% confidence intervals (Cl). Significance levels: * 0.05-0.01, ** <0.01-0.001, ***<0.001.

**Figure 3B:**
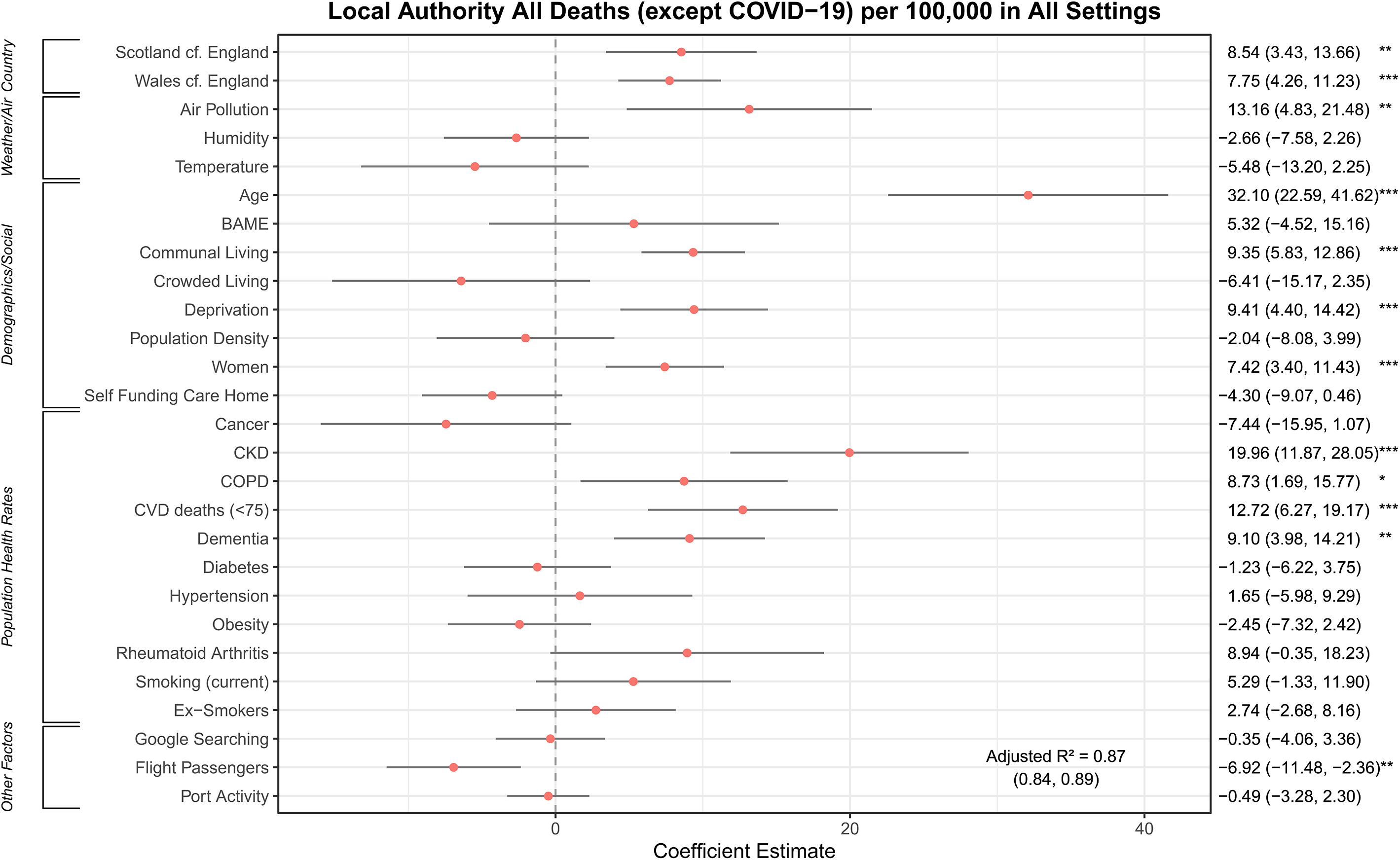
Forest plot of standardised regression coefficients for Local Authority District (LAD) for all deaths per 100,000 of the population except those related to COVID-19 in all settings (care home, hospital and home) with 95% confidence intervals (Cl). Significance levels: * 0.05-0.01, ** <0.01-0.001, ***<0.001.

**Supplementary Table 1** shows the significant results for the same multivariate analyses but the respective deaths in care homes, hospitals and at home separately. These can be interpreted in the context of **Supplementary Figures 2–4**. These data tell us that, relative to COVID-19 unrelated deaths, deaths related to COVID-19 in care homes are predicted by being in Scotland (p = 0.008); however, a higher proportion of ex-smokers (p <0.001) remains protective. Relative to COVID-19 related deaths, COVID-19 unrelated deaths in care homes are associated with higher median age (p = 0.005) and higher rates of communal living (p <0.001), CKD (p = 0.026) and dementia (p <0.001), but being in Wales (p = 0.028) is a protective factor.

Relative to deaths in hospital that are unrelated to COVID-19, COVID-19 related deaths in hospital are higher in LADs with a higher population proportion of BAME origin (p = 0.006) and higher proportion of women (p = 0.006), whereas higher rates of communal living (p = 0.021), self-funding care homes (p = 0.019) and dementia (p = 0.016) are relatively protective. Risk factors for COVID-19 unrelated deaths in hospital (relative to COVID-19 related deaths) are being in Scotland (p = 0.012) and Wales (p <0.001), as well as higher population prevalence of CKD (p = 0.002), COPD (p = 0.004), CVD deaths (p <0.001) and current smoking (p = 0.020). Air pollution is a significant risk factor for both COVID-19 related and COVID-19 unrelated deaths in hospital but is significantly more strongly associated with COVID-19 related deaths (p = 0.015), in-keeping with the results for all combined settings.

Finally, compared with COVID-19 unrelated deaths, COVID-19 death rates at home are higher in LADs with a greater female population proportion (p = 0.023), whereas higher temperatures (p = 0.001) and rates of CVD deaths (p = 0.004) are relatively protective at the LAD level. COVID-19 unrelated deaths at home (relative to COVID-19 deaths) are more strongly associated with humidity (p = 0.020) and higher rates of hypertension (0.025), whereas higher rates of cancer (p = 0.016) and flight passengers (p = 0.013) are relatively protective.

## Discussion

### All Combined Setting Models

In this study of risk factors for COVID-19 related deaths, we have generated explanatory multivariable regression models using the most granular publicly available demographic and ecological data in Great Britain. In contrast with previously published population studies during this pandemic period, we have incorporated both hospital and community death rates across Scotland, England and Wales. Beyond simply identifying significant predictors of COVID-19 related deaths in a multivariable model adjusted for all putative confounders, we have presented this in the context of all GB deaths excluding those related to COVID-19 to identify factors which are significantly unique to COVID-19, thereby minimising over-interpretation of our models and potential collider bias.[55]

From our data, we propose that air pollution (PM2.5 exposure) is the single biggest risk factor for COVID-19 related deaths at a local authority level. In all settings combined, a 1SD increment in population-weighted annual mean PM2.5 (equivalent to 2.00 *μ*g/m3) was associated on average with eight additional deaths per 100,000. A study from the United States similarly looked at PM_2_._5_ exposure, controlling for 20 potential confounding variables, and found that a small increase in longterm fine particulate exposure (1 *μ*g/m^3^) was associated with a significant increase in COVID-19 related death rates (8%).[12] Air pollution causes immune and inflammatory changes both within the lung and systemically, which may be particularly relevant to the excess death rate in COVID-19.[56] Air pollution is also significantly detrimental in the model of COVID-19 unrelated deaths, but the magnitude of the effect is significantly greater in the COVID-19 model **(Table 2)**. Interestingly, our results show that air pollution is significantly positively correlated with higher temperatures **(Figure 2)**. However, a higher temperature is significantly protective against COVID-19 deaths in our models – a factor which has been suggested previously and demonstrated in the first SARS-CoV pandemic.[57]

In spite of consistent evidence showing that being male is a risk factor for in-hospital COVID-19 related deaths, we have shown that, at the LAD level, a higher female population proportion imparts a significantly greater risk. This highlights the importance of careful interpretation of this ecological study: the observation is not related to the rate of female deaths but rather indicates that more deaths overall occur in LAD areas in which more women reside. The underlying relationship between this population-level finding and the observed greater risk of COVID-19 related death among men at the individual level is likely to be complex. A potential explanation might be that women are more likely to be asymptomatic carriers and perpetuate the spread of disease in communities. Indeed, in a study of all women being admitted to an obstetric unit in New York, 88% of all women positive for SARS-CoV-2 were asymptomatic.[58] Women are also more likely to be employed in care-giving roles and involved in direct community infection control measures, such as at testing centres, and this may be a further potential route of transmission.[59]

We observe that a higher rate of ex-smokers in the population is associated with a lower rate of COVID-19 related deaths at the LAD level. Current smoking is not significant in the all combined settings model, but the coefficient is still negative **(Figure 3A)**. While this observation may seem counterintuitive in the context of a respiratory infection, it confirms previous similar findings.[60] Researchers in France have found that smokers were significantly less likely to develop a symptomatic or severe infection from SARS-CoV-2.[61] One possible theory is that nicotine depresses expression of angiotensin-converting enzyme 2 receptors (the functional receptor for SARS-CoV-2)[61], though this effect might be attenuated over time in the case of ex-smokers. Other hypotheses may relate to immunomodulation via nicotinic acetylcholine receptors, leading to decreased production of pro-inflammatory cytokines and chemokines, thereby dampening local inflammation; this mechanism has been suggested in ulcerative colitis where smoking is a known protective factor.[62] The older median age in the LAD is a significant predictor of death only in the model with all deaths excluding those related to COVID-19. Older age has previously been shown to be a significant predictor of in-hospital COVID-19 related deaths.[5,60,63] In view of the aggregate nature of our results, they do not contradict this but rather highlight that, compared to deaths in LADs from all other causes, COVID-19 disease also raises the risk of death in younger people.

While population prevalence of other illnesses (CKD, COPD, CVD deaths and dementia) were significant predictors of COVID-19 unrelated deaths, these were not significantly associated with outcome in the COVID-19 model. Again, this cannot be extrapolated to individual patient risk but does suggest that SARS-CoV-2 related deaths are not patterned according to chronic disease prevalence at the LAD level compared with deaths from other causes. It is also possible that SARS-CoV-2 infection contributed to all-cause deaths, driven by these risk factors, without these deaths being laboratory-confirmed or COVID-19 suspected and entered on the death certificate. Alternatively it may be that individuals with a high burden of disease have experienced excess deaths in the context of reduced general healthcare provision due to scaling back of services during the pandemic and public concerns about accessing healthcare. Future research should focus on ‘excess mortality’ which is the number of deaths recorded during the present pandemic above the level we would have expected under ‘normal’ conditions and would capture all deaths caused by COVID-19, not just those suspected and registered.[64] At the time of analysis, these data was not available at a local authority level.

LADs with higher levels of recorded flight passengers have lower death rates for all deaths except those related to COVID-19. This is correlated with population density and is negatively correlated with median age and so may reflect a younger, more active population. This protective effect is not seen in the COVID-19 model, perhaps due to a detrimental influence of flight activity on population movement and spread in the context of this highly transmissible disease.

Contrary to expectations of a highly contagious disease, population rates of communal living did not significantly associate with COVID-19 related deaths and, if anything, had a negative coefficient. On exploring this further, we note that communal living includes those in protected settings where perhaps strict social distancing measures can be implemented. Further, of the 1.8% of the GB population that reported living in a communal setting, only 31.2% were over the age of 65; the biggest contribution was from 16-24 year olds (39.2%). This group have among the lowest risk of COVID-19 death. Again, there may be an ecological explanation for this apparent anomaly – the highly mobile nature of the 16-24 year old population who are mostly in education may have meant that on the institution of the national lockdown, these groups returned ‘home’ to their local authority of origin where they had resided prior to entering education (e.g. to their parent or guardian’s accommodation). This would deflate the actual population for the LAD in which the communal living establishment was based without altering the denominator population-based on mid-2018 estimates and thus leading to a lowered death rate per 100,000 than would otherwise be expected.

Deprivation prevalence at the LAD level is a risk factor for COVID-19 deaths and COVID-19 unrelated deaths, but there is no statistical difference between the two, suggesting that it is not a factor unique to COVID-19. A higher rate of care home self-funding is strongly negatively correlated with LAD deprivation and, indeed, appears to be protective against COVID-19 related deaths (though, again, this is not significantly different relative to COVID-19 unrelated deaths).

While it has been noted that many people of BAME origin have died from COVID-19 disease, we find that having a higher proportion of BAME residents at the LAD level does not significantly associate with COVID-19 related deaths in the combined settings model. This may reflect the larger urban setting of BAME people and the correlated risks of air pollution and flight passenger numbers, which we accounted for in our analyses.

### Care Home, Hospital and Home Setting Models

Examination of settings separately highlighted setting-specific features. In the care home setting, Scotland LADs were significantly associated with COVID-19 related deaths, compared with those in England. Further investigation may elucidate reasons behind this observation, perhaps related to policy differences on criteria for hospital admissions. Rising death rates in Scottish care homes has recently prompted specific intervention from the Scottish Government.[65] In the care home setting, dementia is a significant risk factor for COVID-19 unrelated deaths only, suggesting that SARS-CoV-2 infection is not dependent on this factor. While not reaching significance in the all combined settings models, LADs with a higher proportion of people of BAME origin have significantly more COVID-19 related hospital deaths, in-keeping with publicised figures.[66,67] As higher proportions of BAME individuals is strongly positively correlated with population density and negatively correlated with higher median age, we hypothesise that younger people living in urban areas are more likely to receive hospital-level care. We also observe that air pollution and a higher proportion of females in the LAD are also strongly associated with hospital COVID-19 deaths. Hospital deaths are the biggest contributor to all COVID-19 related deaths and so these findings are understandably similar to those in the combined settings model. Dementia is a relative protective factor for COVID-19 related hospital deaths, perhaps suggesting that individuals with dementia are less likely to be admitted to hospital. Finally, in a home setting, as with the combined model, LADs with a higher level of flight passengers have lower death rates for COVID-19 unrelated deaths but not for COVID-19 related deaths. Again, this may suggest an influence of movement of young and mobile individuals returning home from other countries or regions on SARS-CoV-2 transmission.

### Limitations

We have reported aggregate data related to local authorities in GB. We acknowledge that such data should not be used to make inferences about individual outcomes (the ecological fallacy). Indeed, one recent study observed that living further from the European Union headquarters was associated with reduced risk of SARS-CoV-2 infection[68]. However, ecological studies do allow us to make inferences about population for public health intervention and are important for hypothesis generation.[69] Further, our study may provide clarity regarding some factors which are only relevant at a local or population level when controlled for societal confounders, such as pollution and climate.[69]

In multivariable models such as ours in which all predictors are modelled together, the interpretation that each predictor is ‘mutually adjusted’ for the others is problematic when some predictors in fact form a mediating pathway towards the outcome. Although we have identified certain risk and protective factors whose association with the outcome is statistically significant even when other covariates are in the model, we have not attempted to conceptualise these in a causal order. Future research using individual-level data should consider plausible hypotheses to be tested in path models, to identify the likely temporal order of risk factors and the optimum point along the path at which an intervention could be targeted.

If data regarding a predictor was available, and measured in the same way at a GB-wide level, this was used. Unfortunately, many of the putative predictor variables were recorded differently and at different times, in the constituent nations of GB. To try to offset these differences, such predictors were z-transformed within the constituent nation prior to their combination across nations. This assumes that the mean and standard deviation of the predictor is similar within each nation. For certain variables, for example deprivation, we know this to be untrue. Deprivation levels have been found to be similar in Scotland and England, but relatively higher in Wales.[70]

Further, occupational risk factors were not included in this analysis. ONS Occupational Coding does not provide sufficient detail to make assumptions about specific occupational exposure and it was not possible to take into account the effects of furlough. Occupation may influence some of our observations – for example, females are more likely to be employed in caring professions which may perpetuate spreading of SARS-CoV-2.

Our study can be compared with that of the OpenSAFELY Group, who studied individual patient data derived from primary care and linked hospital records, facilitating more accurate stratification of patients by past medical history and demographic parameters[6]. While this study benefited from better individual patient granularity, the platform only captures 40% of the population of England and examines in-hospital deaths only. Our study, in contrast, is inclusive of all LADs in England, Scotland and Wales and, uniquely, is informative regarding community death rates.

A number of our references relate to pre-prints and non-academic sources, as is to be expected in the context of this rapidly evolving pandemic. While these need to be interpreted with caution, the wealth of information available is testament to the combined efforts and transparency of our research communities and associated open-access data.

## Conclusions

From this ecological study of risk factors related to COVID-19 related deaths in GB, we have identified significant predictors of death rates in LADs in COVID-19 related cases, in direct comparison with cases not related to COVID-19. We have examined death rates across all hospital and community settings, both combined and separately, and have highlighted factors that appear more relevant in specific settings. We can now hypothesise that air pollution and a higher proportion of females at the local authority level are significantly associated with COVID-19 related deaths. The former is strongly associated and should prompt further studies of individual risk. The latter may relate to asymptomatic carrier status. In contrast to COVID-19 unrelated deaths, COVID-19 related deaths do not seem to be patterned by other illnesses or co-morbidities. Air temperature is protective against COVID-19 deaths at a LAD level. Emergence of data regarding COVID-19 death rate decline in different climates may further elucidate this observation. Ex-smoking appears to protective factor, perhaps related to its modifications on the immune system; clinical trials of immunomodulatory agents are in progress.[71] In specific settings, exploration of reasons behind Scottish care home COVID-19 related deaths and BAME-associated hospital COVID-19 deaths is merited. We anticipate that this comprehensive examination of community and in-hospital data across GB will provide direction for regional and setting-specific research priorities.

#### Summary Box

What is already known on this topic
- The novel SARS-CoV-2 virus has presented Great Britain with an unprecedented challenge in the management and prediction of infection and mortality rates.
- Several individual risk factors for COVID-19 disease mortality have been postulated, including male sex, age, pollution, co-morbidty and ethnicity.
- The influence and interactions of these factors across Great Britain, and in different care settings, is unknown. Relationship of these factors to deaths unrelated to COVID-19 have also yet to be explored.

What this study adds
- In this ecological study of local authority death rates, air pollution was a significant risk factors for mortality from COVID-19 across all care settings. Air temperature, however, was protective.
- Local authority being in Scottish was a significant risk factor for COVID-19 related deaths in the care home setting.
- Local authority having a higher proportion of people from black, Asian and minority ethnic groups was a significant risk factor for COVID-19 related deaths in the hospital setting.
- This study highlights the value of open access data in driving research and informing public policy.

## Data Availability

Data and code are openly available via the link provided.

https://github.com/samleighton87/covid19_LADS/

## Ethics Statement

Ethical approval was not required for this ecological study utilising open data sources.

## Transparency Statement

SPL conceptualised, analysed the data and wrote the manuscript. DJL analysed the data and wrote the manuscript. JH, RU, JC and GG critically evaluated and revised the manuscript. BC and PKM analysed the data and critically evaluated and revised the manuscript. The corresponding author attests that all listed authors meet authorship criteria and that no others meeting the criteria have been omitted. This publication is the work of the authors who will serve as guarantors for the contents of this paper.

